# Computer-aided medical microbiology monitoring tool: a strategy to adapt to the SARS-CoV-2 epidemic and that highlights RT-PCR consistency

**DOI:** 10.1101/2020.07.27.20162123

**Authors:** Linda Mueller, Valentin Scherz, Gilbert Greub, Katia Jaton, Onya Opota

**Affiliations:** Institute of Microbiology, Lausanne University Hospital and University of Lausanne, Lausanne, Switzerland; Infectious Diseases Service, University Hospital of Lausanne, Lausanne, Switzerland

**Keywords:** COVID-19, SARS-CoV-2, RT-PCR, biomedical validation, molecular diagnosis, R-script, algorithm, quality-monitoring, viral load, cycle threshold, clinical evolution, post analytic surveillance

## Abstract

Since the beginning of the COVID-19 pandemic, important health and regulatory decisions relied on SARS-CoV-2 reverse transcription polymerase chain reaction (RT-PCR) results. Our diagnostic laboratory faced a rapid increase in the number of SARS-CoV-2 RT-PCR, with up to 1,007 tests per day. To maintain a rapid turnaround time to support patient management and public health authorities’ decisions, we moved from a case-by-case validation of RT-PCR to an automated validation and immediate transmission of the results to clinicians. To maintain high quality and to track possible aberrant results, we developed a quality-monitoring tool based on a homemade algorithm coded in R.

We present the results of this quality-monitoring tool applied to 35,137 RT-PCR results corresponding to 30,198 patients. Patients tested several times led to 4,939 pairwise comparisons; 88% concordant and 12% discrepant. Among the 573 discrepancies, 428 were automatically solved by the algorithm. The most likely explanation for these 573 discrepancies was related for 44.9% of the situations to “Clinical evolution”, 27.9% to “Preanalytical” problems, and 25.3% to “Stochastic”. Finally, 11 discrepant results could not be explained, including 8 received from external partners for which clinical data were not available.

The implemented quality-monitoring strategy allowed to: i) assist the investigation of discrepant results ii) focus the attention of medical microbiologists onto results requiring a specific expertise and iii) maintain an acceptable TAT. This work highlighted the high RT-PCR consistency for the detection of SARS-CoV-2 and the importance of automated processes to handle a huge number of samples while preserving quality.

## Introduction

The rapid spread and the high incidence of COVID-19 pandemic caused unprecedented challenges for diagnostic microbiology laboratories. Rapid and high throughput SARS-CoV-2 reverse transcription polymerase chain reaction (RT-PCR) have been quickly developed as the ones proposed by Corman et al. (1-3). These molecular diagnostic assays rapidly became the cornerstone of patient diagnosis as well as hospital and public health managements. Consequently, microbiology laboratories were reorganized to respond to the high demand for SARS-CoV-2 testing (4). This situation required (i) the rapid adaptation of infrastructures, (ii) quick validation and implementation of new RT-PCR assays, (iii) working hour extension and the employment of new workforces. Yet, the overall quality in SARS-CoV-2 results as well as for all other results provided by clinical microbiology laboratories had to be maintained throughout the crisis.

Validation procedures had to be simplified in response to the very high demand for SARS-CoV-2 testing. Our molecular diagnostic laboratory located in a tertiary care university hospital faced a rapid increase in the number of SARS-CoV-2 PCR with up to 1,007 tests per days at the peak of the epidemic. To ensure the best quality, two validation steps are usually applied prior results transmission to clinicians: the technical validation of the assay by laboratory technicians is followed by the biomedical validation of the results by medical microbiologists, who consider the specific clinical setting (5). However, biomedical validation quickly appeared as a bottleneck in the SARS-CoV-2 analytical workflow since it extends the turnaround time (TAT), at the risk of affecting clinical outcomes as well as infection prevention strategies and adequacy of public health decisions (6). Thus, to maintain an acceptable TAT, results were released to the clinicians after technical validation based on the FastFinder software (UgenTec NV, Hasselt, Belgium) that automatically analyses RT-PCR amplification curves.

The limited experience on these newly implemented RT-PCR assays, including their performance (7), highlighted the need for an active surveillance of the quality of provided results. Delta checks are commonly used in clinical chemistry laboratories to monitor analytical throughputs that outreach capacity for sample-by-sample validation. Delta checks describes a process where discrepancies in sequential results of the same patient are detected to prompt repetition of the analysis (8). We wondered whether a similar approach could be used to monitor the quality of SARS-CoV-2 results obtained in our laboratory. However, Delta checks are usually restricted to analytes exhibiting limited short-term variations and is as such unsuitable to microbiology results. Thus, we developed a quality monitoring methodology based on a homemade algorithm programmed in R to monitor SARS-CoV-2 RT-PCR results. The developed quality monitoring methodology leveraged repeated testing to identify potential preanalytical or analytical culprits as well as cases requiring further biomedical investigations. The algorithm developed in-house accounts for the expended variability among results, to restrict the list of discrepancies to cases truly requiring investigation.

In this article, we present the results obtained from the application of our quality surveillance on data from the first four months of SARS-CoV-2 crisis in our laboratory. Beside its role as quality management tool, application of this surveillance allowed us to quickly gain knowledge about this RT-PCR applied to a novel virus and new disease. In particular, this process allowed us to identify clinical specimen with significant added value and the presence of long-term carrying patients.

## Materials and Methods

### RT-PCR and samples

Samples collected from patients with a suspected COVID-19 or for screening were tested by RT-PCR, using either our high-throughput MDx platform (5), the cobas SARS-CoV-2 qualitative test (Roche, Basel, Switzerland) and the Xpert SARS-CoV-2 test (Cepheid, California, USA). The E gene was targeted by the RT-PCR performed on the MDx platform (5), as described by Corman and colleagues (1). The cobas SARS-CoV-2 targeted the E gene as well as the ORF1/a and was performed according to the manufacturer guidelines. Finally, the Xpert SARS-CoV-2 test targets the N gene and the E gene. The three methods displayed similar performances for the detection of SARS-CoV-2 from various clinical specimens and similar cycle threshold (Ct) value when positive (9-11). Samples were mainly collected from the upper respiratory tract. However, other type of samples were also tested (Table S1). Nasopharyngeal and oropharyngeal swabs were collected in Copan Universal Transport Medium System (UTM-RT) or BD™ Universal Viral Transport System (UVT). Sputa and bronchial aspirates were liquefied using N-acetyl-L-cysteine prior analysis using the cobas 6800 system, or prior nucleic acid extraction using the MagNAPure 96 instrument when samples are tested on our MDx Platform (12, 13). Anorectal swabs were collected as previously described (14).

### Data collection and analysis

Data were collected during the first four months of the epidemic in Switzerland (12.02.2020-12.06.2020) and included all SARS-CoV-2 RT-PCR analyses conducted at the Institute of Microbiology of the Lausanne University Hospital (CHUV). SARS-CoV-2 RT-PCR results and basic contextual information were extracted from our Laboratory Information System (LIS) (MOLIS, CGM) and analyzed with the R (15)(version 3.6.1) programming language helped by packages from the *Tidyverse (16)* environment.

### Discrepant cases identification and classification

A R script was developed in-house to automatically identify and classify discrepant cases. In this script, all analyses from patients with multiple samples were compared to their previous results in a pairwise approach (https://github.com/valscherz/SARS-CoV-2_discrepant_screen). Samples comparisons were then categorized as concordant or discrepant. Only discrepant results were further processed. These discrepancies (positive versus negative or conversely) between consecutive samples were classified based on i) Ct values, ii) samples types and iii) reception dates (Fig. S1). Based on these records, discrepancies were classified by the algorithm as following:

- “Stochastic” classification was retained by the algorithm when the positive sample of the discrepant pair presented a Ct values over 35; close to the analytic limit of detection of the method; a Ct of 35 corresponds to 3,800 copies/ml).
- “Low yield” described samples type rarely or never observed as positive (i.e. less than 5% of times, Table S1). Thus, the algorithm classified as “Low yield” discrepancies where a low yield sample returned a negative result (explainable by sample nature, e.g. blood and urine).
- “Time delay” explanation was retained by the algorithm when the time interval between the two compared samples was over 10 days. Indeed, in this case, the discrepancy would be explained by the evolution of the disease (new infection or disease resolution).
- “To be investigated” was finally retained by the algorithm for discrepancies meeting none of these criteria and requiring further investigation by medical microbiologists.

The script was designed to compare each sample only to the last relevant result. For instance, a positive nasopharyngeal swab followed by a negative PCR in blood, and then later by another positive nasopharyngeal swab, will lead to only one discrepancy: the negative blood classified as a “Low yield”. The second nasopharyngeal swab will be classified as concordant with the previous nasopharyngeal swab. Of note, two nasopharyngeal samples taken more than 10 days apart once negative and once positive with a Ct > 35, would be classified as “Stochastic” and not as “Time delay”. Indeed, discrepancies were classified according to the first matching criteria in the following order: “Low yield”, “Stochastic”, “Time delay” and “To be investigated”, as detailed in the graphical scheme of the decisional algorithm (Fig. S1). Furthermore, a result from a patient with 3 samples or more can be involved both in a concordant and a discrepant pairwise comparisons. Indeed, the second of his analyses could be in agreement with the first of his results but discrepant with the third.

Discrepant samples classified as “To be investigated” by this algorithm were then manually curated, classified and assigned to a reason putatively explaining the discrepancy (Table S3). In this manual analysis, discrepancies between two nasopharyngeal swabs taken within the same period (< 24h) and collected in different units or different hospitals (compatible with differences in sampling quality) were manually classified as imputable to “Sample quality”. When sampling sources were different (i.e. comparing an upper respiratory tract sample with a rectal swab), samples were manually classified as “Different sample types”. Discrepancies between samples collected less than 10 days apart and with indications in clinical records supporting a recent infection or recent recovery were classified as “Clinical context”. Finally, discrepancies compatible with none of these putative explanations were classified as “Unsolved”. For visualization purposes, classified discrepancies were regrouped into corresponding testing phases or context: clinical context, preanalytical or stochastic (Tables S2 and S3).

## Results

### Included SARS-CoV-2 RT-PCR results

Since the implementation of SARS-CoV-2 RT-PCR assays and for a period of four months, 30,198 patients were tested by RT-PCR at the Institute of Microbiology of the Lausanne University Hospital (CHUV). This corresponded to 35,137 samples, among which 4,545 (12.9%) returned a positive result, whereas 30,592 (87.1%) were negative. Upper respiratory tract (URT) samples represented 98% of the tested specimen (Table S1). The peak of number of analyses took place on March 18^th^ with up to 1,007 analyses processed on the same day (Fig. 1).

**Figure 1.**
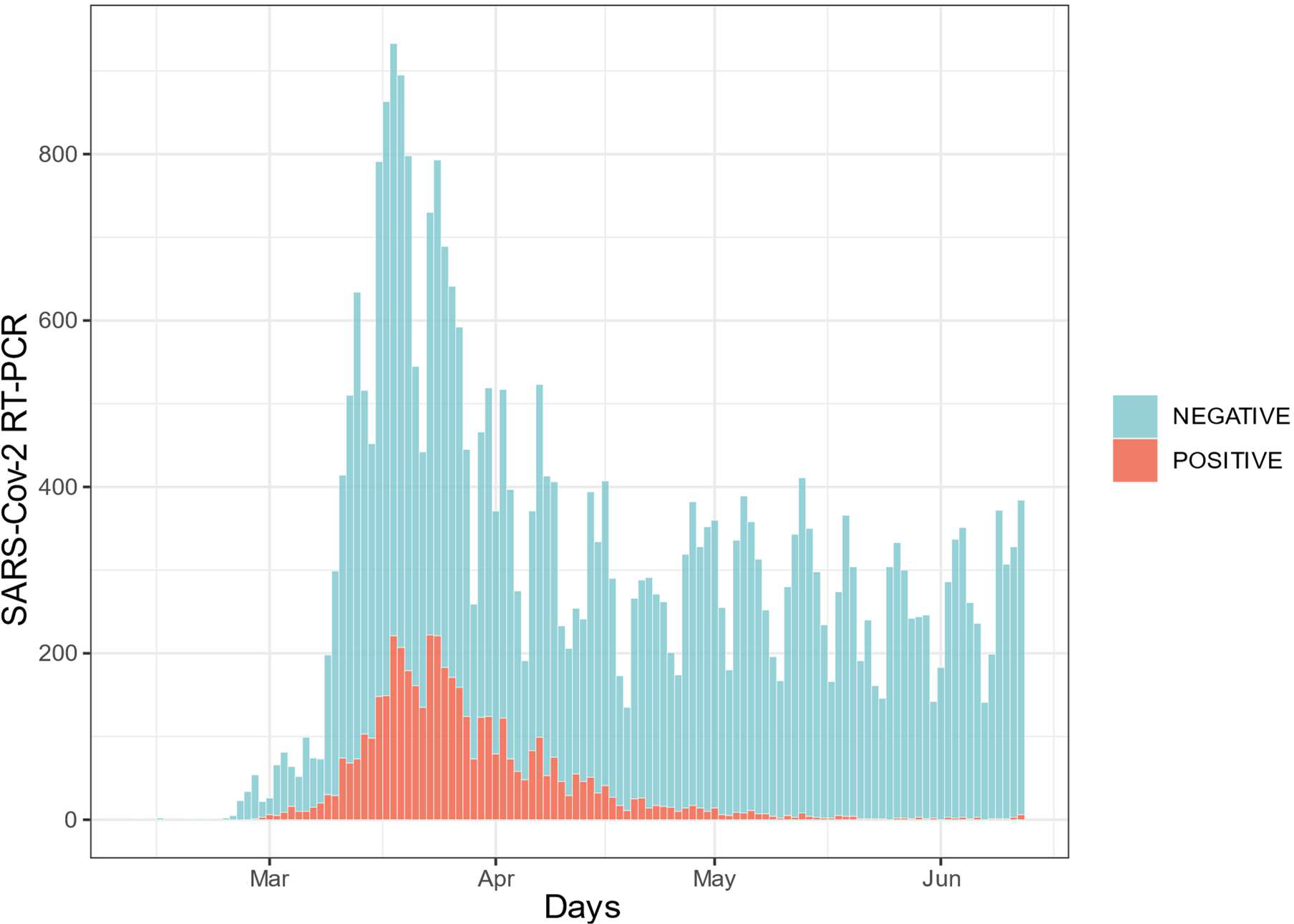
Daily number of SARS-CoV-2 qPCR assays. 35,137 analyses are represented here. Data are distributed according to their reception date. Blue bars represent samples for which RT-PCR results were negative (88%) while orange bars depict positive samples (12%).

### Post-analytic surveillance of RT-PCR results by an in-house informatic algorithm

Due to the difficulty to maintain a systematic sample-by-sample biomedical validation of such a high number of tests, systematic biomedical validation of all results was suspended and a surveillance supported by an algorithm encoded in R language was developed. This algorithm aimed at supporting the identification and investigation of discrepancies for repeated testing, as well as gaining insight on quality of the analyses provided by our laboratory. Applied to the first four months of SARS-CoV-2 testing in our laboratory, the designed algorithm identified 2,792 patients with only concordant results, either negative (2,704) or positive (88), among the 3,214 patients owing at least two specimens (Fig. 2). Conversely, 422 patients presented at least one discrepant pair of results. Of note, 127 patients had concordant as well as results; these latter were further processed by the algorithm as patients with discrepant results. When considering 4,939 pairwise comparisons of successive results for these patients with multiple tests, 4,366 (88%) were concordant and 573 (12%) were discrepant. Among these concordant pairwise comparisons, 250 (6%) were positive results while 4,116 were negatives (94%) (Fig. 2).

**Figure 2.**
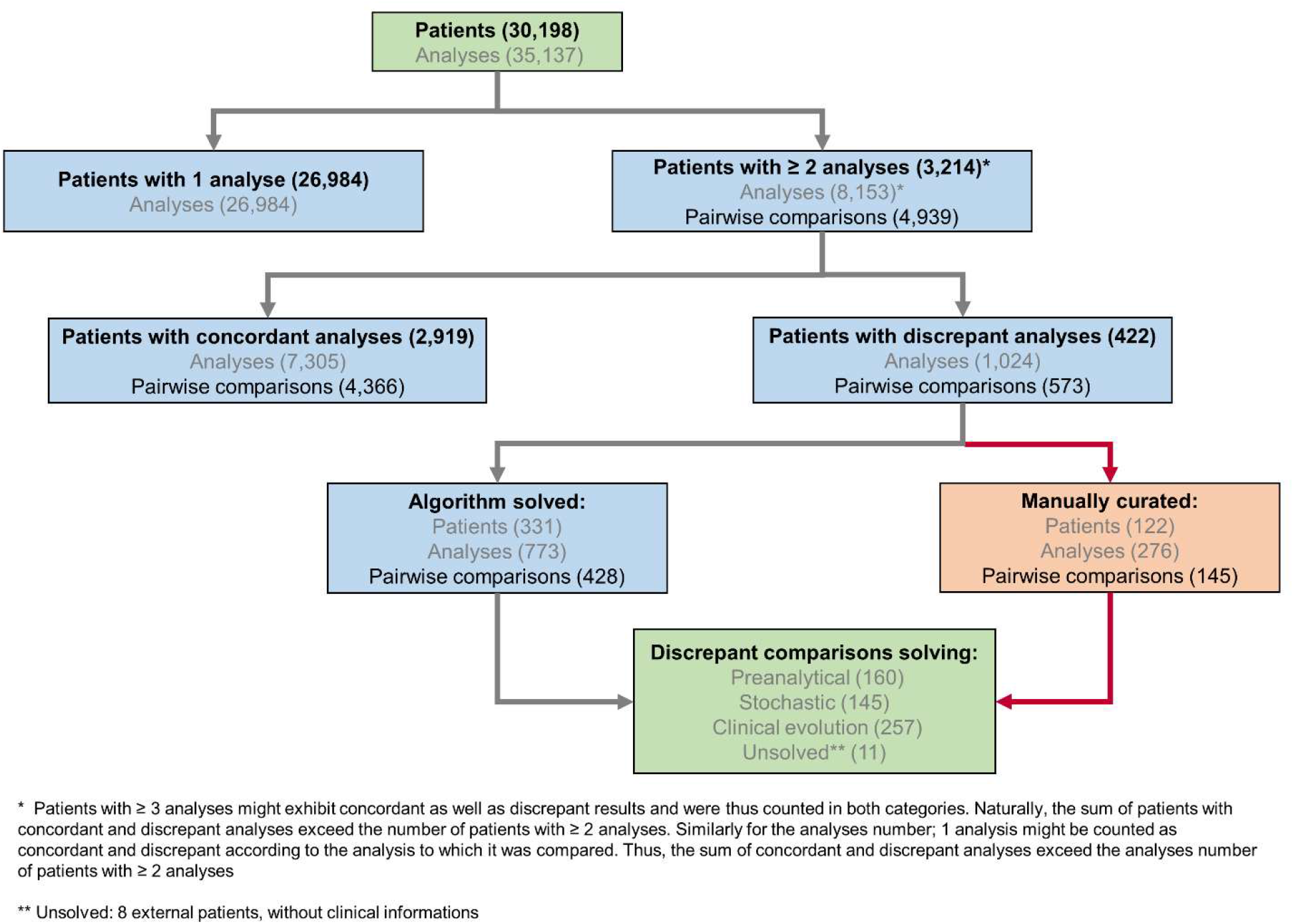
SARS-CoV-2 analytical flowchart. An R-based script was used to identify patients with multiple, potentially discrepant, results (upper part of the chart) and therefore insure their surveillance. The algorithm processed further, discrepant results and a potential explanation for the observed discrepancy was attributed either automatically either manually (lower part of the chart and Fig. S1)

### Algorithm-based and manual biomedical investigation of discrepancies

Our pipeline significantly reduced the number of discrepancies requiring human investigation and a probable explanation could be identified for most of the discrepant results. Indeed, the majority of the discrepant pairwise comparisons (n=428/573) could be automatically attributed by the pipeline to a putative explanation (“stochastic”, “low yield” or “time delay”) (Fig. 3A and 3B, Table S2). Only the remaining discrepancies (n=145) did not fit any of the solving rules encoded in the algorithm and required investigations based on the available analytical and clinical information (Table S3).

**Figure 3.**
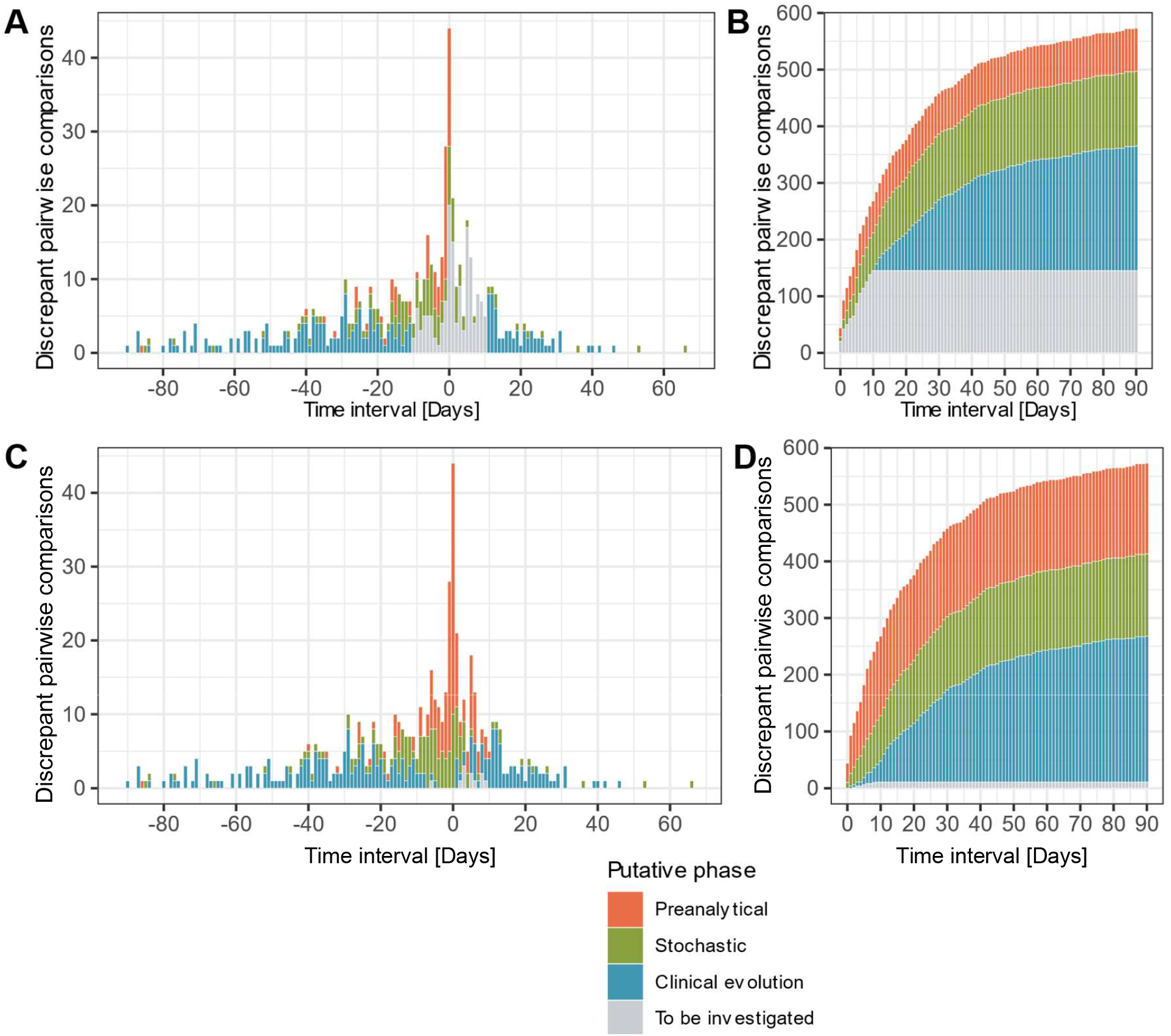
Time laps between discrepant analyses, putative phase assignment and cumulative score. 573 pairwise comparisons are represented according to their date of reception and colored depending on the analytical phase which best explained the observed discrepancy. **A and B**. Before manual curation discrepancies were classified by the algorithm as “Preanalytical”, “Stochastic”, “Clinical evolution” and “To be investigated”. **C and D**. After manual curation comparisons previously categorized as “To be investigated” were re-assigned to the same categories or as “Unsolved”. **A and C**. Time laps between discrepant analyses. Transitions from a negative to a positive result are represented in the positive side of × axe, while positive to negative transitions are plotted on the negative side. **A**. Before manual investigation. Only comparisons of samples taken less than 10 days apart required investigation since the “Clinical evolution” was otherwise assigned by default by the algorithm. **C**. After manual investigation. Short term discrepancies were mostly attributed to “Preanalytical” factors and “Stochastic” phenomenon. Discrepant analyses putatively explained by “Clinical evolution” of the disease was naturally retained for samples collected within larger time interval. **B and D**. Cumulative score of discrepant analyses. **B**. 74.7% of comparisons were solved and classified, while 25.3% were classified as “To be solved” and needed further investigations. **D**. In a 90 days interval cumulative score shows that most discrepancies could be explained by the “Clinical evolution” of the disease. Details on discrepancies putative classification and phases are detailed in Materials and Methods section.

The profiles of putative explanations for discrepancies evolved depending on the time interval between the compared analyses (Fig. 3C). In samples received among the same day, our assessment explained 77.3% (n=34/44) of the discrepancies as related to the preanalytical phase (i.e. explained by the sample types or sample collection in different health centers), followed by stochasticity (Ct value >35) at 22.7% (n=10/44). The discrepancies in results of samples received 1-3 days apart were explained by factors affecting the preanalytical phase at 55.4% (n=51/92), followed stochasticity at 32.6% (n=30/92). Interestingly, 7 of the discrepancies observed in the 1-3 days interval were explainable by nosocomial (n=6) or community (n=1) acquired infections based on health records, which could explain the quick negative to positive transition. These 7 discrepancies were thus classified in the clinical evolution context. As for the 4 remaining discrepancies, clinical records were not available for 3 and the last one remained unexplained. Investigations of discrepancies between samples received 4-10 days apart incriminated the preanalytical phase at 41.6% (n=55/132), followed by stochasticity at 30.3% (n=40/132). As expected, the discrepancies imputable to the clinical evolution of the disease based on clinical records (new infection or infection resolution) was greater in the 4-10 days interval since it represented 22.7% of the discrepancies (n=30/132). The 7 remaining discrepancies in this time interval could not be explained, either in absence (n=5) or in presence (n=2) of clinical information. Over 10 day, the clinical evolution of the disease was the main explanation (72.1%, n=220/305) for discrepancies, as it was the default explanation retained by our automatic pipeline for discrepant results from samples collected more than 10 days apart in absence of any other explanation.

In the overall assessment of the 573 discrepancies from samples taken 90 days apart, 44.9% (n=257) of discrepancies could be explained by the clinical evolution of the disease (e.g. indications in clinical records for new contagion, time delay making new infection or infection resolution likely) (Fig 3D). 27.2% (n=160) of cases had arguments for factors incriminating the preanalytical phase (discrepant results among samples collected by different health centers, inclusion of samples rarely positive as blood); and 25.3% (n=145) of the discrepant comparisons could be explained by analytical stochasticity in presence of low RNA loads (Ct value > 35 for the positive sample followed or preceded by a negative sample). No clear explanation could be identified for 1.9% (n=11) of the discrepancies (classified as “Unsolved”). For 8 of the unsolved situations, samples were submitted to our laboratory by external care centers or private laboratories and clinical records were thus not accessible. No explanation for discrepancies could be found for 3 cases, despite the availability of full clinical documentation. Moreover, short-term negative to positive transitions were compatible with 21 nosocomial and 8 community-acquired infections based on clinical records (Table S3).

### Evolution of the discrepancy patterns across the epidemic period

The pattern in transitions (negative result followed by a positive result or the reverse) among discrepancies evolved over the studied period of four months. In the first two months included in the analysis, which corresponds to the first two months of the epidemic in our region (12.02-12.04.2020), 71.3% of negative to positive transitions were observed (n=154/216). Conversely, in the last two months (13.04-12.06.2020), 81.2% on the transitions went from a positive to a negative result (n=290/357), which contributed to the overall trend of 61.4% of positive to negative discrepancies (n = 352/573) (Figs. 3A and 3C). Such observation was expected and imputable to follow-up of resolving infections.

### Sustained RT-PCR positive results in patients

Besides discrepancies, clinicians also investigated sustained positivity in patients. In our analysis, the longest time interval between two positive results from the same patient was of 83 days (Figs. 4B and 5, please note that Fig. 4B describes the time interval between successive concordant positive analyses from the same patient while Fig. 5 considers the time interval from the first to the last positive sample of patient identified as long-time carriers). Considering the time interval between their first and the last positive result, we observed 32, 11, and 3 patients with sustained positivity in samples taken over 30, 50 and 70 days apart, respectively. This observation questions the presence of active viral replication or only of viral traces left from the resolving infection. Indeed, the last sample of these long time carriers displayed a low viral loads with Ct values around 35 (corresponding to 3,800 copies/ml) in all but one notable exception (Fig. S2).

**Figure 4.**
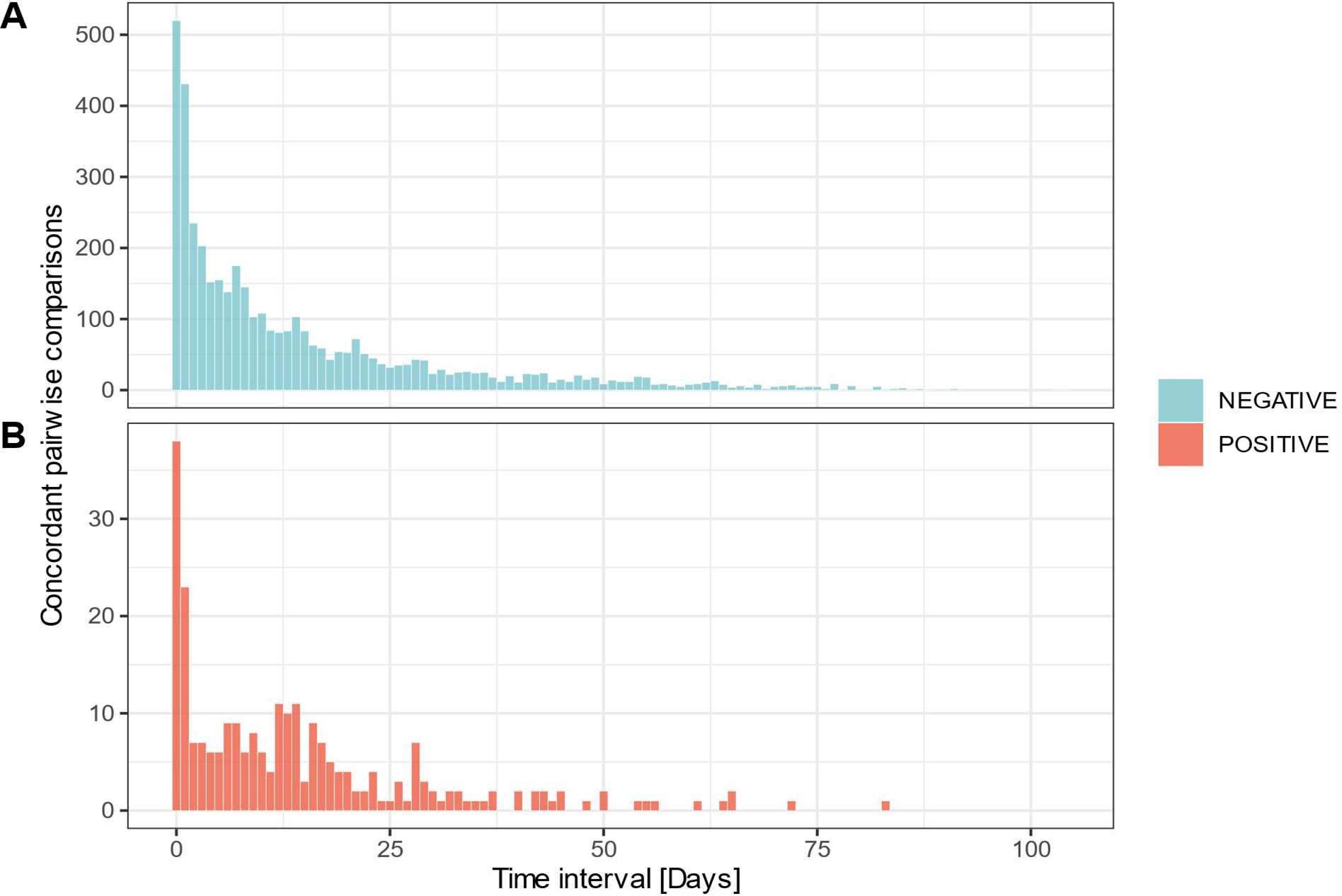
Time laps between concordant analyses. Distribution of 4,366 pairwise concordant comparisons according to the sample reception date. **A**. Concordantly negative comparisons for SARS-CoV-2 detection (n=4,116). **B**. Concordantly positive comparisons for SARS-CoV-2 detection (n=250).

**Figure 5.**
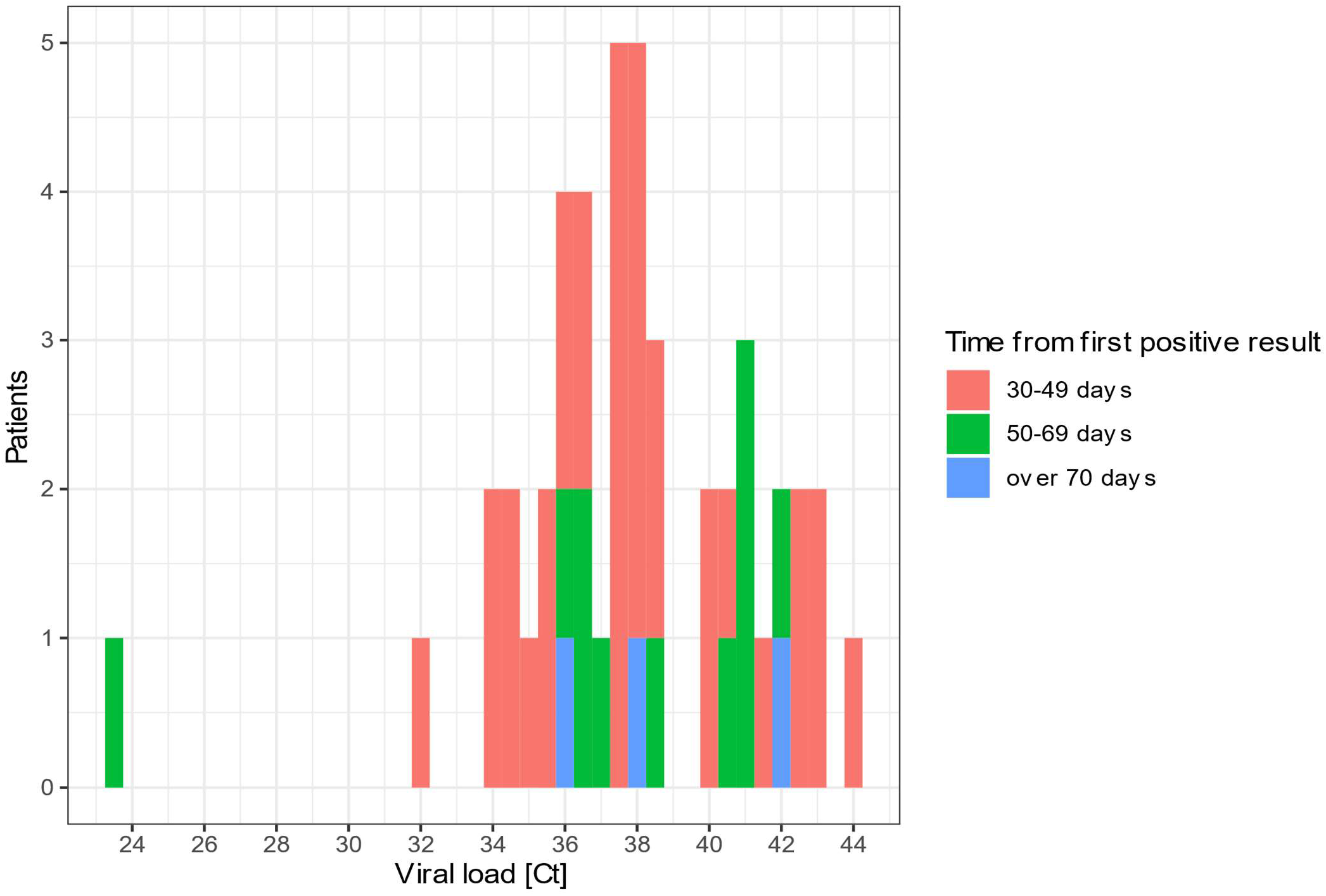
Distribution of Ct values for patients with sustained RT-PCR positive results. This histogram represents the Ct value (maximal if more than 1) for SARS-CoV-2 detection by RT-PCR in the last positive sample of patients with sustained positivity in samples taken 30 days apart or more.

### Pairwise analyses highlight the consistency of SARS-CoV-2 RT-PCR results

Reports questioned early in the performance of RT-PCR for SARS-CoV-2 detection, which supported recommendations for repeated testing (17). Yet, in our dataset only 2.5% of the negative results obtained from an URT sample (n=733/29,714) were followed by an additional analysis in between 1h and 3 days after initial testing. In comparison, 0.6% of cases (28/4,451) of the positive results from URT samples were followed by a second analysis over the same time interval. Thus, if repeated testing remained limited, a negative result was still significantly more often challenged than a positive result by clinicians (Pearson’s Chi-squared Test, p < 0.001, OR = 4.0).

An evaluation of the performance of the RT-PCR for SARS-CoV-2 detections based on samples collected over a short time interval showed a good level of negative and positive agreements. Indeed, an initial negative result on an URT sample was confirmed in 99.2% (243/245) of cases for patients tested twice on the same day (Table 1); both discrepancies could be explained by stochasticity since associated to high Ct values. Conversely, a first positive result was confirmed in 88.9% (24/27) of cases; two discrepancies could be explained by stochasticity too, while the third involved a positive nasopharyngeal swab and a negative throat swab, a sample site shown to be less sensitive for SARS-CoV-2 detection (18). As expected, agreement rates diminished with the time: concordance negative rate for URT samples switched from 99.2% for samples collected along the same day to 94.2% for samples collected in between 1-3 days. Respectively, the concordance positive agreement passed from 88.9% to 70.0%.

**Table 1.**

| URT samples | Same day | 1-3 day | 4-6 days | 7-10 days |
| --- | --- | --- | --- | --- |
|  | n = 272 | n = 826 | n = 489 | n = 585 |
| Concordant Negative results | 243 | 750 | 430 | 516 |
| Concordant Positive results | 24 | 21 | 7 | 20 |
| Negative to Positive transition | 2 | 46 | 38 | 28 |
| Positive to Negative transition | 3 | 9 | 14 | 21 |
| **Concordant negative agreement** | 99.2% | 94.2% | 91.9% | 94.9% |
| **Concordant positive agreement** | 88.9% | 70% | 33.3% | 48.8% |

## Discussion

This work presents the outcome of a homemade algorithm developed in response to the need for quality surveillance of SARS-CoV-2 RT-PCRs which throughput exceeded the ability to conduct manual biomedical validation for each sample. Applied to the 35,137 SARS-CoV-2 RT-PCRs performed in our diagnostic laboratory from February 12 to June 12, 2020, the algorithm identified 3,214 patients owing multiple tests. These patients represented an opportunity for quality assessment of our analyses, but also required careful attention to investigate potential discrepancies. Among the 3,124 patients tested multiple times, we observed, supported by our in-house R algorithm, a majority (86.8%) of concordant results, mostly negative (96.8%). Conversely, the algorithm identified 422 patients with at least one pair of discrepant results. The algorithm solved 74.7% of the discrepant results thus dramatically reducing the number of cases (145) requiring biomedical investigations. Together, the clinical evolution of the disease (44.9%), preanalytical factors (27.9%) and stochasticity around the limit of detection (25.3%) were the most likely explanations retained by the automatic algorithm-based classification and the biomedical validation for the 573 observed discrepancies. Only 1.4% of the cases remained unexplained because clinical records were not available. Despite availability of all records, 0.5% of the results remained unexplained.

The natural evolution of the disease appeared as the main origin for discrepancies among repeated testing in our classification. Indeed the clinical context contributed to explain almost half (44.9%) of the discrepancies assessed by automatic resolution and manual investigation together. The positive to negative transition, compatible with a solved infection, was the most frequent pattern (61%). This is consistent with the studied period, which integrated the development of the epidemic in our region and the transition phase where fewer cases occurred and the infection resolved in a majority of patients. The automatic algorithm classified all discrepancies between samples collected more than 10 days apart as related to the clinical evolution of the disease, in absence of any other putative explanation. This rule was designed to limit the need for manual review of discrepant cases that were not expected to be informative, but could have biased discrepancies classification toward this category.

Preanalytical factors were incriminated in 27.9% of the observed discrepancies. Negative result from samples types rarely or never positive, such as blood and urine, was the most frequent explanation retained for these discrepancies (47.5%) (Table S2). Among the remaining 52.5% of discrepancies associated to preanalytical phase, 25.0% were due to the comparisons of two samples of different nature (i.e. an URT sample and a rectal swab). Both of these specimens presented over 5% of positivity in our dataset and thus were considered as samples with an interesting clinical yield. Yet, positivity in only one of the sampling site has already been reported (19). Thus, comparisons of distinct sample collection sites was considered in our assessment as a sufficient explanation for the observed discrepancy. The remaining 27.5% of the discrepancies, occurred after repeated collection of samples in less than 10 days but in different healthcare provider (Table S2). While only hypothetical, this classification suggests that the performance of sampling might rely on sampling technique and other preanalytical factors such as storage time and sampling conditions. Altogether, these preanalytical factors and the expected inconsistency of viral shedding (20) in nasopharyngeal mucosa could contribute to explain the sub-optimal performance of nasopharyngeal swabs for SARS-CoV-2 detection reported in recent studies (7, 21). Furthermore, the attribution of most of the observed discrepancies to the preanalytical phase was in agreement with previous reports that described this testing phase as most frequent source of errors in general in clinical laboratories (22, 23).

Stochasticity of the RT-PCR reaction, explaining 25.3% of the observed discrepancies, was the only source of disagreeing results directly related to the analytical phase. Stochasticity - here estimated at a Ct of 35 corresponding to 3,800 copies/ml - is expected in viral load near to the limit of detection. The clinical impact of these discrepancies could be limited since low viral loads are expected in the late course of the disease and when the infectivity might be diminished (10, 11, 24).

Based on repeated testing, our results support the good performance of RT-PCR in URT samples for SARS-CoV-2 detection. Former publications questioned the performance of such analyses (7, 25), and encouraged repeated testing by clinicians. The positive and negative agreements of RT-PCR results among URT samples collected the same day were of 88.9% and 99.2% respectively. The positive concordance rate exhibited an important drop when samples were collected more than 1 day apart, with the lowest concordance agreement (33.3%) observed in a 4-6 days time delay. This might be explained by the clinical evolution of patients. Nevertheless, these positive agreement rates should be considered with caution, due to the small number of positive samples for which additional testing were requested (i.e. in a time interval of 4-6 days 468 negative samples were retested, while for positive samples only 21 were reanalyzed). In fact, patients were 4x times more often retested 1-72 hours after a first initial negative results (2.5%) than after a first positive result (0.6%) offering more opportunities to assess negative than positive agreement.

A limitation of our quality surveillance methodology is that it leaves aside patients tested only once. However, repeated testing was recommended for results in disagreement with the clinical context. Thus, one can assume that most of the results of patients tested only once met clinical expectations, supporting the adequacy of the provided analyses. Furthermore, results quality was insured by a technical validation. Such validation relies on: i) positive and negative controls integrated to each run, ii) the use of two RT-PCR targets and iii) the per-batch follow-up of the positivity rate; if a run would suddenly have a much higher rate of positivity would suggest a possible contamination problem with false positive. Additionally, the follow-up of the median quantity of virus was also used to identify a possible technical issue affecting the sensitivity. However, our work might be complemented by a systematic, unbiased repetition of analyses for quality purposes. Nevertheless, in the crisis we faced, the upfront repetition of analyses would have been impractical, and maybe even unethical, in a context of reagents and material shortage.

Another limitation of our work is that while we intended to use an unbiased algorithm stable over time to investigate discrepancies in results, some of the applied criteria were partly arbitrary (e.g. the 10 days limit to consider discrepancies as expected due to the clinical evolution of the disease). Furthermore, our process consent the use of a single explanation for each observed discrepancy, while more could be applicable. While arguable, these choices were made to fit a strategy of quality monitoring. Indeed, the primary aim of the presented methodology was to attribute the observed discrepancies to the most likely explanation to focus on truly unexplainable and problematic cases.

Clinical laboratory vulnerabilities during the COVID-19 pandemic were the subject of a recent publication by Lippi *et al*. (26). Our assessment overlaps with some of the preanalytical culprits identified by the authors such as specimen collection (see “detailed explanations” Tables S2 and S3). However, some other potential vulnerabilities were not considered as probable causes for discrepancies in our assessment, since they are covered by other pre-existing quality management procedures in our laboratory. For instance, our preanalytical team systematically rejects samples missing a patient identification. Moreover, in our laboratory, internal extraction controls and amplification controls are systematically included to detect samples that might contain interfering substances compromising the amplification (9).

Repeated testing on identical samples is a recognized method for internal quality assurance, which can lead to the identification of systematic or sporadic vulnerabilities in microbiology laboratories (27). In the present quality monitoring, different samples from the same patient were compared, similarly to the “Delta check” routinely applied in clinical chemistry laboratories. However, microbiology laboratories present specific challenges due to the frequent occurrence of actual discrepant results drew by clinical evolution of the diseases, different sampling sites, robustness of the sampling procedure or stochasticity in presence of scarce DNA. These factors were considered by our monitoring tool to reduce the list of potentially problematic cases to 3 discrepant comparisons. Unfortunately, these 3 suspicious cases remained unaddressed at the time of this final analysis. If considered as erroneous results, 3 over the 35,349 analyses provided on the period of four months would be an acceptable error rate (0.01%), especially when compared to previous reports on error rates (0.1-9.3%) in medical laboratories data (28). Furthermore, similar cases would now, and thanks to our algorithm implemented in routine, lead to repetition of the analyses or feedback to the clinicians to obtain new samples.

This is, to our knowledge, the first implementation in clinical microbiology of a quality monitor tool, similar to the “Delta check” used in clinical chemistry laboratories. We would recommend the implementation of a similar system for other laboratories facing the COVID-19 or other epidemic wave in order to help focusing on some specific results and to identify laboratory vulnerabilities. Furthermore, we hypothesized that similar longitudinal observation of results and algorithm-based selection of “cases to investigate” could also be applied to other high-throughput microbiology laboratory assays, either by the implementation of ad-hoc software as presented here or by rules embedded in the LIS.

This work emphasized the benefit of an automatic algorithm capable of finding discrepant results and attributing them to corresponding testing phases. This computer-aided methodology outlined that besides the expected evolution of the disease, most of discrepant results are compatible with preanalytical factors. Moreover, most of URT samples collected repeatedly in a short timeframe showed consistent results, displaying the good reproducibility of the RT-PCR for SARS-CoV-2 detection. Application of this method for quality monitoring enabled to focus on problematic cases requiring biomedical expertise while maintaining an acceptable TAT.

## Data Availability

All data referred to in the manuscript are available

## Acknowledgements

We would like to deeply thank all the staff of the Institute of Microbiology of the Lausanne University Hospital. In particular, we thank all the staff of the Laboratory of Molecular Diagnostic of the Institute of Microbiology of the University of Lausanne and all the training FAMH of the COVID-19 post-analytic team of our institute.

## References

1. Corman VM, Landt O, Kaiser M, Molenkamp R, Meijer A, Chu DK, Bleicker T, Brunink S, Schneider J, Schmidt ML, Mulders DG, Haagmans BL, van der Veer B, van den Brink S, Wijsman L, Goderski G, Romette JL, Ellis J, Zambon M, Peiris M, Goossens H, Reusken C, Koopmans MP, Drosten C. 2020. Detection of 2019 novel coronavirus (2019-nCoV) by real-time RT-PCR. Euro Surveill 25.

2. Tadini EP-O, M. Opota, O. Moulin, E. Lamoth, F. Manuel, O. Lhopitallier, L. Jaton, K. Croxatto, A. Grandbastien, B. Senn, L. Guery, B. 2020. SARS-CoV-2, un point dans la tourmente. Rev Med Suisse 16:917–23.

3. Caruana G, Croxatto A, Coste AT, Opota O, Lamoth F, Jaton K, Greub G. 2020. Diagnostic strategies for SARS-CoV-2 infection and interpretation of microbiological results. Clin Microbiol Infect doi:10.1016/j.cmi.2020.06.019.

4. Posteraro B, Marchetti S, Romano L, Santangelo R, Morandotti GA, Sanguinetti M, Cattani P, Group FCL. 2020. Clinical microbiology laboratory adaptation to COVID-19 emergency: experience at a large teaching hospital in Rome, Italy. Clin Microbiol Infect doi:10.1016/j.cmi.2020.04.016.

5. Greub G, Sahli R.,Brouillet, R., Jaton, K. 2015. Ten years of R&D anf full automation in molecular diagnosis. Future Microbiology 11:403–425.

6. Hawkins R. 2007. Laboratory Turnaround Time. Clin Biochem Rev 28:179–194.

7. Kokkinakis IS, K. Favrat, B. Genton, B. Cornuz J., 2020. Performance du frottis nasopharyngé-PCR pour le diagnostic du Covid-19 Recommandations pratiques sur la base des premières données scientifiques. Rev Med Suisse 16.

8. Schifman RB, Talbert M, Souers RJ. 2017. Delta Check Practices and Outcomes: A Q-Probes Study Involving 49 Health Care Facilities and 6541 Delta Check Alerts. Arch Pathol Lab Med 141:813–823.

9. Poljak M, Korva M, Knap Gasper N, Fujs Komlos K, Sagadin M, Ursic T, Avsic Zupanc T, Petrovec M. 2020. Clinical evaluation of the cobas SARS-CoV-2 test and a diagnostic platform switch during 48 hours in the midst of the COVID-19 pandemic. J Clin Microbiol doi:10.1128/JCM.00599-20.

10. Moraz M, Jacot D, Papadimitriou-Olivgeris M, Senn L, Greub G, Jaton K, Opota O. 2020. Clinical importance of reporting SARS-CoV-2 viral loads across the 2 different stages of the COVID-19 pandemic. MedRXiV.

11. Jacot D, Greub G, Jaton K, Opota K. 2020. Viral load of SARS-CoV-2 across patients and compared to other respiratory viruses. MedRXiV.

12. Opota O, Brouillet R, Greub G, Jaton K. 2017. Methods for Real-Time PCR-Based Diagnosis of Chlamydia pneumoniae, Chlamydia psittaci, and Chlamydia abortus Infections in an Opened Molecular Diagnostic Platform. Methods Mol Biol 1616:171–181.

13. Opota O, Zakham, F., Mazza-Stalder, J., Nicod, L., Greub, G., Jaton, K.. 2019. Added Value of Xpert MTB/RIF Ultra for Diagnosis of Pulmonary Tuberculosis in a Low-Prevalence Setting. J Clin Microbiol 57:e01717–18.

14. Dang T, Jaton-Ogay K, Flepp M, Kovari H, Evison JM, Fehr J, Schmid P, Boffi El Amari E, Cavassini M, Odorico M, Tarr PE, Greub G. 2009. High prevalence of anorectal chlamydial infection in HIV-infected men who have sex with men in Switzerland. Clin Infect Dis 49:1532–5.

15. Team RC. 2019. R: A language and environment for statistical computing. R Foundation for Statistical Computing, Vienna, Austria.

16. Wickham H, Averick M, Bryan J, Chang W, McGowan L, François R, Grolemund G, Hayes A, Henry L, Hester J, Kuhn M, Pedersen T, Miller E, Bache S, Müller K, Ooms J, Robinson D, Seidel D, Spinu V, Takahashi K, Vaughan D, Wilke C, Woo K, Yutani H. 2019. Welcome to the Tidyverse. Journal of Open Source Software 4.

17. Patel R, Babady E, Theel ES, Storch GA, Pinsky BA, St George K, Smith TC, Bertuzzi S. 2020. Report from the American Society for Microbiology COVID-19 International Summit, 23 March 2020: Value of Diagnostic Testing for SARS-CoV-2/COVID-19. mBio 11.

18. Wang X, Tan L, Wang X, Liu W, Lu Y, Cheng L, Sun Z. 2020. Comparison of nasopharyngeal and oropharyngeal swabs for SARS-CoV-2 detection in 353 patients received tests with both specimens simultaneously. Int J Infect Dis 94:107–109.

19. Chen Chen GG, Yanli Xu,Lin Pu, Qi Wang, Liming Wang,. 2020. SARS-CoV-2– Positive Sputum and Feces After Conversion of Pharyngeal Samples in Patients With COVID-19. Annals of Internal Medicine doi:10.7326/M20-0991.

20. Qi L, Yang Y, Jiang D, Tu C, Wan L, Chen X, Li Z. 2020. Factors associated with duration of viral shedding in adults with COVID-19 outside of Wuhan, China: A retrospective cohort study. Int J Infect Dis doi:10.1016/j.ijid.2020.05.045.

21. Yang Y, Yang M, Shen C, Wang F, Yuan J, Li J, Zhang M, Wang Z, Xing L, Wei J, Peng L, Wong G, Zheng H, Liao M, Feng K, Li J, Yang Q, Zhao J, Zhang Z, Liu L, Liu Y. 2020. Evaluating the accuracy of different respiratory specimens in the laboratory diagnosis and monitoring the viral shedding of 2019-nCoV infections. MedRXiV doi:10.1101/2020.02.11.20021493.

22. Plebani M. 2006. Errors in clinical laboratories or errors in laboratory medicine? Clin Chem Lab Med 44:750–9.

23. West J, Atherton J, Costelloe SJ, Pourmahram G, Stretton A, Cornes M. 2017. Preanalytical errors in medical laboratories: a review of the available methodologies of data collection and analysis. Ann Clin Biochem 54:14–19.

24. Yu F, Yan L, Wang N, Yang S, Wang L, Tang Y, Gao G, Wang S, Ma C, Xie R, Wang F, Tan C, Zhu L, Guo Y, Zhang F. 2020. Quantitative Detection and Viral Load Analysis of SARS-CoV-2 in Infected Patients. Clin Infect Dis doi:10.1093/cid/ciaa345.

25. Williams TC, Wastnedge E, McAllister G, Bhatia R, Cuschieri K, Kefala K, Hamilton FJ, Johannessen I, Laurenson IF, Shepherd J, Stewart A, Waters D, Wise H, Templeton K. 2020. Sensitivity of RT-PCR testing of upper respiratory tract samples for SARS-CoV-2 in hospitalised patients: a retrospective cohort study. MedRXiV doi:10.1101/2020.06.19.20135756.

26. Lippi G, Plebani M. 2020. The critical role of laboratory medicine during coronavirus disease 2019 (COVID-19) and other viral outbreaks. Clin Chem Lab Med doi:10.1515/cclm-2020-0240.

27. Scherz V, Durussel C, Greub G. 2017. Internal quality assurance in diagnostic microbiology: A simple approach for insightful data. PLoS One 12:e0187263.

28. Kalra J. 2004. Medical errors: impact on clinical laboratories and other critical areas. Clin Biochem 37:1052–62.

